# Cyclodialysis Surgery for Enhanced Uveoscleral Outflow and Intra-Ocular Pressure Lowering in Glaucoma: A Systematic Review and Meta-Analysis of 100 Years of Clinical Evidence

**DOI:** 10.1101/2025.04.05.25325239

**Authors:** Robert Stamper, Alex Huang, Carol Toris, Mary Qiu, Gerry Gray, Reena Garg, Tsontcho Ianchulev

## Abstract

**Objective:** To perform a systematic review and meta-analysis of the clinical evidence of the treatment effect of surgical cyclodialysis in the management of intraocular (IOP) in patients with glaucoma.

**Methods:** A comprehensive literature review was conducted of peer-reviewed interventional studies from the PubMed, Cochrane, Web of Science and EMBASE databases of surgical cyclodialysis treatment for the lowering of intraocular pressure in patients with glaucoma. Key outcome measures of treatment success were long-term IOP control, as well as IOP-lowering medication burden and the incidence of intraoperative and postoperative adverse events. The meta-analysis was registered with Prospero ID CRD42025632759

**Results:** A total of 40 studies spanning a publication period of more than 100 years of surgical cyclodialysis treatment encompassing data from over 4,082 eyes were included in the analysis. Clinical evidence comprised observational, non-randomized studies, 75% of which involved an ab-externo approach and 25% comprised an ab-interno cyclodialysis intervention. Given the natural evolution of the clinical paradigm over the years, changes in surgical technique, instrumentation and addressable population, the overall analysis was constructed to account for the significant variability in outcomes reporting. Across the final evaluable dataset, the clinical performance of cyclodialysis surgery was characterized by overall qualified success rates of 72.3% on average (range 33%-97%) over a postoperative follow-up period ranging from 6 to 132 months. Depending on surgical technique and disease severity, reported success rates indicate slightly increased efficacy and lower rate of complications with ab-interno intervention. Durability of the cyclodialysis procedure varied significantly, with higher rates of failure in patients with advanced and refractory glaucoma. Specific complications such as persistent hyphema, hypotony and vision loss were reported infrequently. All outcomes, including IOP reduction, ocular safety, and durability, showed significant improvement with the newer interventional ab-interno surgical techniques.

**Conclusion:** Cyclodialysis remains an enduring surgical intervention and one of the few available surgical options for uveoscleral outflow enhancement in glaucoma patients. The IOP lowering effect of the procedure can be significant, albeit variable, with better clinical performance in mild and moderate glaucoma and with advanced interventional ab-interno surgical approaches.

**Synopsis:** Results of a meta-analysis comprising more than 4,000 glaucoma cases of cyclodialysis surgery for the lowering of intraocular pressure demonstrate significant and sustained efficacy of one of the few surgical interventions for uveoscleral outflow enhancement.

## INTRODUCTION

Glaucoma represents the leading cause of irreversible blindness globally, affecting over 80 million individuals ^1^. Treatment options are aimed at lowering of the Intraocular Pressure (IOP) starting with medications or laser trabeculoplasty ^2,3^ as a first-line treatment, followed by surgical intervention if pharmacotherapy fails. The major objective of surgical glaucoma management involves enhancing aqueous humor drainage, either through ab-interno procedures targeting the trabecular or uveoscleral outflow pathways, or externally via transscleral filtration techniques, such as trabeculectomy and glaucoma drainage devices (GDDs) ^4,5,6^.

Cyclodialysis, first introduced by Leopold Heine in 1905 enhances uveoscleral outflow. It was one of the most prevalent surgical treatments until the 1960’s before the advent of trabeculectomy and glaucoma drainage implants. The more recent innovations of ab-interno suprachoroidal stents and the emergence of MIGS have spurred renewed interest in cyclodialysis intervention a minimally invasive access point to uveoscleral space ^7,8^. This technique has had over a century of clinical practice with variable acceptance. Mechanistically, cyclodialysis involves the creation of an internal aqueous outflow channel, facilitating egress through the uveoscleral pathway via the suprachoroidal and trans-scleral routes ^9^. The goal of cyclodialysis is to augment the native uveoscleral outflow pathway through an iatrogenically created cleft that acts as an iatrogenic aqueous conduit and ab-interno filtration reservoir. Cyclodialysis clefts appear to play a role in the natural physiology of uveoscleral outflow. Small endogenous ciliary body clefts have been documented in histological sections and during ocular patient imaging ^10,11,12,13,14^. These clefts are felt to be important for uveoscleral outflow as their presence or absence correlates with higher or lower uveoscleral outflow in both animals and humans ^15,16,17^. The creation of a surgical cyclodialysis is intended to further augment the uveoscleral outflow capacity of the endogenous clefts for the lowering of IOP.

The uveoscleral pathway provides a potentially advantageous surgical target due to its function as a low-resistance sink for aqueous drainage. From studies of the pharmacologic treatment of glaucoma, selective agents for uveoscleral outflow, such as prostaglandin analogues, have been shown to have a higher therapeutic index for aqueous drainage enhancement compared to trabecular outflow drugs. The intrinsic pressure gradient between the anterior chamber and the supraciliary space, coupled with the absorptive properties of the choroid, generates highly favorable conditions for internal aqueous outflow ^18^. The osmotic gradient across the choriocapillaris is essential in promoting fluid movement in response to the hydrostatic pressure differential between the anterior chamber and the suprachoroidal space ^19.20,21^. Notably, while uveoscleral outflow typically functions as a pressure-independent mechanism, the introduction of a cyclodialysis cleft reduces ciliary muscle resistance, rendering the outflow pressure-dependent ^22,23^. Physiological studies indicate that uveoscleral outflow accounts for approximately 40% to 50% of aqueous drainage in humans and non-human primates, with higher rates observed in pediatric populations, which gradually decrease with age ^24,25,26^.

The surgical technique for the creation of an iatrogenic cyclodialysis has undergone significant refinements over the past century. Heine’s original ab-externo approach, known as the classical cyclodialysis, involved a limbus-based concentric ab-externo scleral incision positioned 5 mm posterior to the limbus. A spatulated instrument was then used to disinsert the ciliary body and create the cleft. Over time, advancements in surgical instrumentation facilitated a shift towards a more interventional ab-interno technique in order to minimize tissue trauma and better surgical control and visualization of the cyclodialysis construction. More recent modifications include various strategies to reinforce the filtration channel to increase durability of the internal filtration channel and reduce the risk of closure and restenosis. These reinforcement techniques involve the use of air, viscoelastic substances, scleral allografts, and other scaffolding materials to maintain patency of the cyclodialysis cleft.

This systematic review consolidates over a century of clinical evidence from the peer-review literature on the efficacy and safety of cyclodialysis surgery for IOP reduction in the treatment of glaucoma.

## METHODS

### Search Strategy and Study Selection

A comprehensive literature search encompassing the PubMed, Cochrane, Web of Science, and EMBASE databases was conducted to identify studies published between 1905 and 2024. The following search terms were applied: “glaucoma” “open angle glaucoma” “closed angle glaucoma” “mixed mechanism glaucoma” “secondary glaucoma” “congenital glaucoma” (population), “cyclodialysis” “ab-interno cyclodialysis” “ab-externo cyclodialysis” (intervention), and “intraocular pressure” (outcome). Synonymous terms were combined using the Boolean operator “OR” while the primary search terms were interconnected using “AND” An additional search was performed excluding the outcome as a restrictive term to capture descriptive studies, particularly those published prior to the 1950s, which often lacked standardized outcome measures. The search encompassed all glaucoma populations. Titles, abstracts, and full texts were meticulously screened to ensure consistency with analytical specifications.

No additional restrictive terms were used for any of the databases and a broad inclusion criteria were maintained to minimize the risk of bias. Extracted outcomes were systematically tabulated from each study. Due to the heterogeneity and variability of outcome measures reported over a century of publications, multiple outcomes were extracted where available. The most consistently reported outcome across historic and contemporary studies was the percentage of patients achieving post-operative success in terms of “qualified” intraocular pressure (IOP) control.

While there was significant variability in the authors’ definitions of successful IOP control across the studies, in the absence of access to primary patient results for each study, we used the proportion of study subjects achieving qualified post-operative success (with or without use of adjunct IOP lowering medications), as reported by the authors, as our main efficacy outcome measure. A subset of recent studies reported mean changes in medicated IOP from baseline, and these measures were incorporated where available. Outcomes were extracted at the latest follow-up time points for each study. Reported incidences of procedure-related ocular adverse events were also evaluated.

### Analysis Methods

Meta-analytic methods were used to summarize qualified success outcomes from the 40 included studies. Procedure types were defined and analyzed by group: ab-externo, ab-interno, ab-interno with sub-scleral reinforcement of the cleft, stand-alone procedures and procedures in combination with other surgery (e.g. cataract surgery). Due to smaller numbers some subgroupings could not be statistically characterized.

The base model was a random-effects meta-analysis, with study as the random effect, assuming that each study has a true qualified success rate and that these follow a normal distribution. Studies were weighted relative to sample size. An additional fixed effect was added to the base model to investigate the effect of each of: procedure type, approach, stand-alone versus combination procedures, year published, and follow-up time. Each fixed effect was evaluated in a separate analysis using a mixed effects model with study as the random effect. Estimates, standard errors, and confidence intervals are calculated based on the fitted random effects or mixed effects model. Fitting was carried out using R version 4.3.0 and the R package for version 4.6-0. REML methods were used in the fitting. The meta-analysis was registered with PROSPERO ID CRD42025632759.

## RESULTS

### Study Selection and Data Extraction

In the EMBASE database, 828 research articles met the initial search criteria for cyclodialysis, while 202 references met the criteria in PubMed as of 12/14/2024. The initial search of Web of Science yielded 164 citations. A systematic review applying additional restriction criteria focused on treatment-specific cyclodialysis in glaucoma patients, thereby excluding studies of traumatic or iatrogenic cyclodialysis, suprachoroidal aqueous drainage devices, epidemiologic studies, veterinary reports, and non-surgical investigations. The dataset was further enriched by including pre-1950 publications referenced in the reviewed literature. Ultimately, 40 studies encompassing 4,082 surgical cases were included in the final analysis. The steps and the results of the PRISMA procedure are summarized in Figure 1.

**Fig 1.**
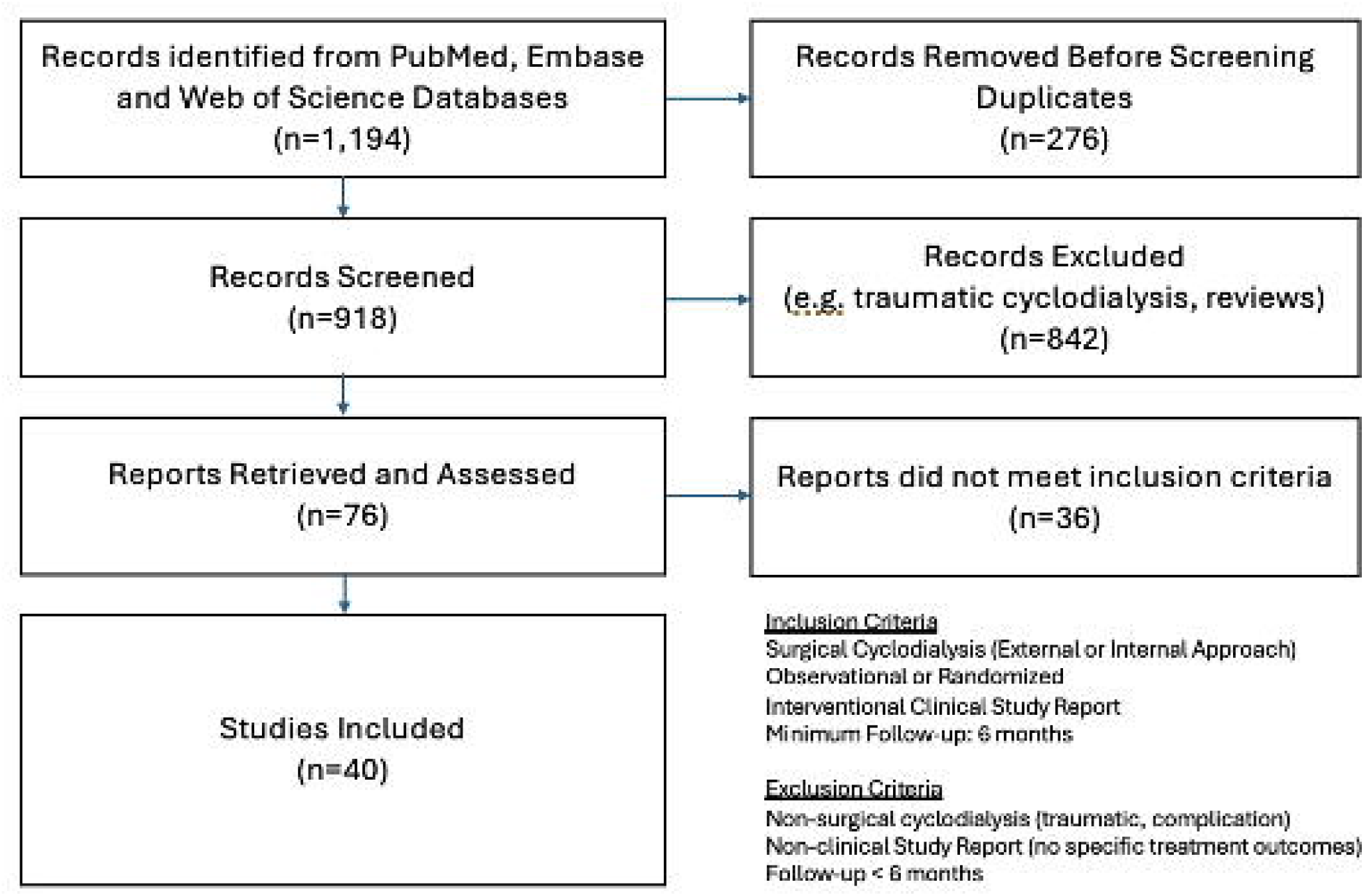
PRISMA Flowchart

All the studies were further assessed and categorized for the level of bias using the ROBINS-I v2 systematic tool (https://sites.google.com/site/riskofbiastool/welcome/robins-i-v2). Given the non-randomized, uncontrolled, observational nature of the peer-review dataset, there were no studies in the low-risk category. All studies were in the moderate-high risk categories.

Follow-up durations ranged from 6 to 132 months, and the patient cohort spanned a wide patient age group - from a pediatric population (<1 year) to elderly patients (>90 years). The studies included a diverse range of glaucoma subtypes, such as primary and secondary open-angle glaucoma as well as angle-closure glaucoma. Cyclodialysis techniques varied across studies, with earlier publications focusing on standard ab-externo procedures and more recent studies emphasizing ab-interno approaches. Several studies incorporated adjunct cataract procedures and sub-scleral reinforcements to enhance the durability and efficacy of cyclodialysis surgery. Table 1 summarizes the characteristics of the included studies.

**Table 1.**
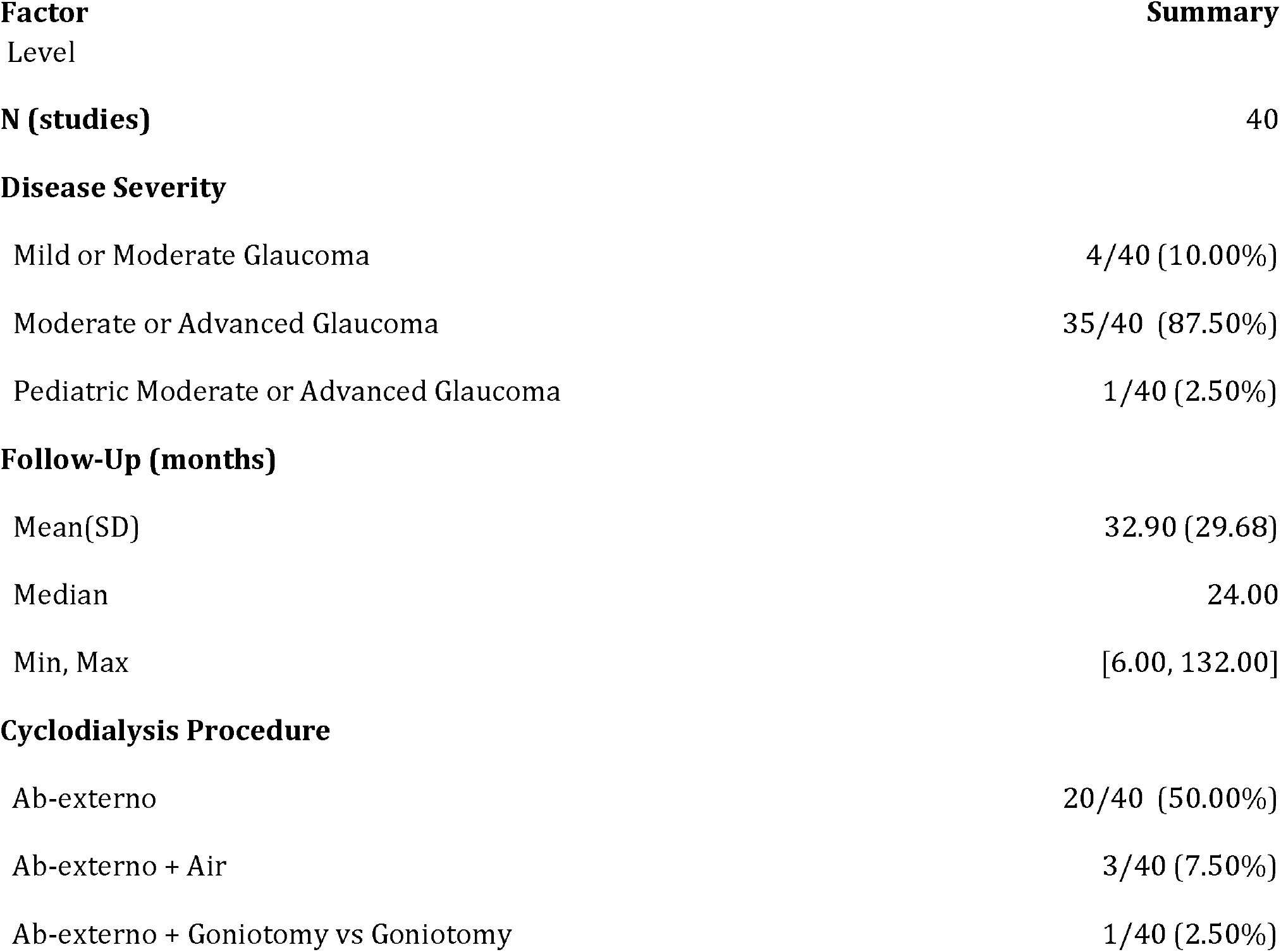
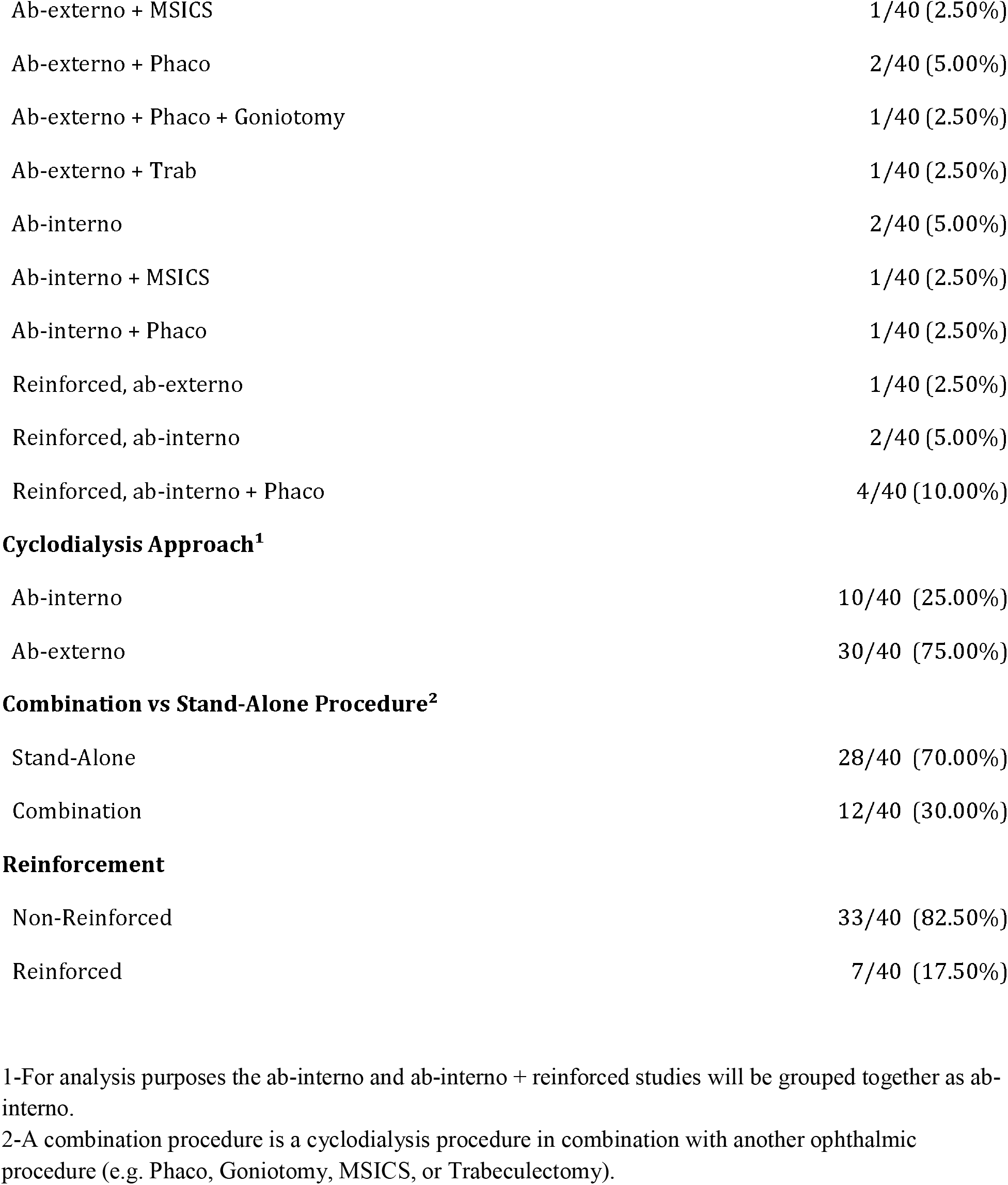
Summary of Study Characteristics.

| **Factor** | **Summary** |
| --- | --- |
| **Level** |  |
| **N (studies)** | 40 |
| **Disease Severity** |  |
| Mild or Moderate Glaucoma | 4/40 (10.00%) |
| Moderate or Advanced Glaucoma | 35/40 (87.50%) |
| Pediatric Moderate or Advanced Glaucoma | 1/40 (2.50%) |
| **Follow-Up (months)** |  |
| Mean(SD) | 32.90 (29.68) |
| Median | 24.00 |
| Min, Max | [6.00, 132.00] |
| **Cyclodialysis Procedure** |  |
| Ab-externo | 20/40 (50.00%) |
| Ab-externo + Air | 3/40 (7.50%) |
| Ab-externo + Goniotomy vs Goniotomy | 1/40 (2.50%) |
| Ab-externo + MSICS | 1/40 (2.50%) |
| Ab-externo + Phaco | 2/40 (5.00%) |
| Ab-externo + Phaco + Goniotomy | 1/40 (2.50%) |
| Ab-externo + Trab | 1/40 (2.50%) |
| Ab-interno | 2/40 (5.00%) |
| Ab-interno + MSICS | 1/40 (2.50%) |
| Ab-interno + Phaco | 1/40 (2.50%) |
| Reinforced, ab-externo | 1/40 (2.50%) |
| Reinforced, ab-interno | 2/40 (5.00%) |
| Reinforced, ab-interno + Phaco | 4/40 (10.00%) |
| **Cyclodialysis Approach<sup>1</sup>** |  |
| Ab-interno | 10/40 (25.00%) |
| Ab-externo | 30/40 (75.00%) |
| **Combination vs Stand-Alone Procedure<sup>2</sup>** |  |
| Stand-Alone | 28/40 (70.00%) |
| Combination | 12/40 (30.00%) |
| **Reinforcement** |  |
| Non-Reinforced | 33/40 (82.50%) |
| Reinforced | 7/40 (17.50%) |

1-For analysis purposes the ab-interno and ab-interno + reinforced studies will be grouped together as ab-interno.
2-A combination procedure is a cyclodialysis procedure in combination with another ophthalmic procedure (e.g. Phaco, Goniotomy, MSICS, or Trabeculectomy).

### Summary of Systematic Analysis

All included studies were observational, comprising both retrospective and prospective designs; no randomized controlled trials (RCTs) were identified in the peer-reviewed literature. Among the 40 studies involving 4,082 patients, 30 studies (3,183 patients) evaluated cyclodialysis performed through an external surgical approach (ab-externo), while 10 studies (899 patients) assessed interventional cyclodialysis using an internal approach (ab-interno).

### Meta-Analysis of Outcomes

The estimated overall qualified success rate from the meta-analysis across all studies was 72.7%, with a 95% CI of [67.7%, 77.6%]. Outcomes were similar regardless of Procedure Type, Approach, or Stand-Alone versus Combination procedures. Formal statistical tests of the fixed effects for procedure type, approach, and stand-alone vs combination were not significant. (Table 2, Figure 2)

**Table 2.** Meta-Analysis of Qualified Success Rates: Overall, with fixed effect for Procedure Type, with fixed effect for Approach, and with fixed effect for Stand-Alone/Combination.

| **Meta-Analysis** | **n (studies)** | **Estimate (SE)** | **95% CI** |
| --- | --- | --- | --- |
| **All** |  |  |  |
| All | 40 | 72.7% (2.4%) | [67.9%, 77.6%] |
| **Approach** |  |  |  |
| Externo | 28 | 77.5% (4.9%) | [67.6%, 87.4%] |
| Interno | 12 | 71.2% (2.7%) | [65.7%, 76.8%] |
| **Stand-Alone vs Combo** |  |  |  |
| Stand-Alone | 10 | 71.4% (2.9%) | [65.6%, 77.2%] |
| Combination | 30 | 75.8% (4.4%) | [66.9%, 84.7%] |
| **Reinforcement** |  |  |  |
| Non-Reinforced | 33 | 70.9% (2.6%) | [65.7%, 76.0%] |
| Reinforced | 7 | 81.9% (5.6%) | [70.5%, 93.3%] |
| **Follow-Up Time** |  |  |  |
| <=20 months | 17 | 74.6% (3.7%) | [67.1%, 82.1%] |
| 21-40 months | 11 | 75.3% (4.7%) | [65.8%, 84.9%] |
| 40-80 months | 9 | 70.2% (4.9%) | [60.2%, 80.1%] |
| >80 months | 2 | 54.8% (10.5%) | [33.4%, 76.2%] |

Meta-analytic estimates are based on a random-effects meta-analysis with study as the random effect. For Procedure Type, for Approach, and for Stand-Alone vs Combo, a fixed effect was added to the model. For the Procedure Type analysis all ab-interno procedure types, with or without reinforcement are combined into one category. For the Stand-Alone vs Combo analysis, a combination procedure is defined as a cyclodialysis procedure in combination with another ocular surgery with an IOP lowering effect (e.g. cataract surgery)

**Table 3.** Final list of studies included in the evidence-based review of cyclodialysis surgical treatment.

| Study No. | Year Pub. | Study Type | Disease Severity | Cyclodialysis Procedure | Study Follow-up (months) | N | Qualified Success Rate |
| --- | --- | --- | --- | --- | --- | --- | --- |
| 1 | 1920 | Observational | Moderate or Advanced Glaucoma | Ab-externo | 12 | 350 | 0.72 |
| 2 | 1920 | Observational | Moderate or Advanced Glaucoma | Ab-externo | 24 | 37 | 0.81 |
| 3 | 1930 | Observational | Moderate or Advanced Glaucoma | Ab-externo | 60 | 120 | 0.72 |
| 4 | 1931 | Observational | Moderate or Advanced Glaucoma | Ab-externo | 24 | 692 | 0.78 |
| 5 | 1937 | Observational | Moderate or Advanced Glaucoma | Ab-externo | 12 | 36 | 0.33 |
| 6 | 1940 | Observational | Moderate or Advanced Glaucoma | Ab-externo | 12 | 51 | 0.93 |
| 7 | 1941 | Observational | Moderate or Advanced Glaucoma | Ab-externo | 60 | 121 | 0.523 |
| 8 | 1946 | Observational | Moderate or Advanced Glaucoma | Ab-externo | 120 | 206 | 0.52 |
| 9 | 1946 | Observational | Moderate or Advanced Glaucoma | Ab-externo | 36 | 140 | 0.42 |
| 10 | 1947 | Observational | Moderate or Advanced Glaucoma | Ab-externo + Air | NA | 83 | 0.75 |
| 11 | 1947 | Observational | Moderate or Advanced Glaucoma | Ab-externo | 60 | 121 | 0.471 |
| 12 | 1947 | Observational | Moderate or Advanced Glaucoma | Ab-externo + Air | 12 | 57 | 0.877 |
| 13 | 1949 | Observational | Moderate or Advanced Glaucoma | Ab-externo | 24 | 100 | 0.79 |
| 14 | 1949 | Observational | Moderate or Advanced Glaucoma | Ab-externo | 60 | 100 | 0.79 |
| 15 | 1954 | Observational | Moderate or Advanced Glaucoma | Ab-externo | 24 | 11 | 0.727 |
| 16 | 1957 | Observational | Moderate or Advanced Glaucoma | Ab-externo | 60 | 153 | 0.85 |
| 17 | 1957 | Observational | Moderate or Advanced Glaucoma | Ab-externo | 60 | 102 | 0.774 |
| 18 | 1958 | Observational | Moderate or Advanced Glaucoma | Ab-externo | 15 | 90 | 0.767 |
| 19 | 1958 | Observational | Moderate or Advanced Glaucoma | Ab-externo | 24 | 22 | 0.727 |
| 20 | 1960 | Observational | Moderate or Advanced Glaucoma | Ab-externo | 60 | 60 | 0.535 |
| 21 | 1969 | Observational | Moderate or Advanced Glaucoma | Ab-externo + Phaco | 72 | 48 | 0.893 |
| 22 | 1976 | Observational | Moderate or Advanced Glaucoma | Ab-externo | 12 | 78 | 0.87 |
| 23 | 1986 | Observational | Moderate or Advanced Glaucoma | Ab-interno + Phaco | 15 | 16 | 0.938 |
| 24 | 1986 | Observational | Moderate or Advanced Glaucoma | Ab-externo | 15 | 16 | 0.625 |
| 25 | 1995 | Observational Pediatric | Pediatric Moderate or Advanced Glaucoma | Ab-externo + Phaco + Goniotomy | 12 | 98 | 0.77 |
| 26 | 1998 | Observational | Moderate or Advanced Glaucoma | Ab-externo + MSICS | 29 | 50 | 0.74 |
| 27 | 2013 | Observational | Moderate or Advanced Glaucoma | Ab-externo + Air | 12 | 32 | 0.75 |
| 28 | 2013 | Observational | Moderate or Advanced Glaucoma | Ab-externo + Phaco | 60 | 50 | 0.74 |
| 29 | 2015 | Observational | Moderate or Advanced Glaucoma | Ab-externo + Trab | 132 | 55 | 0.58 |
| 30 | 2019 | Observational | Moderate or Advanced Glaucoma | Reinforced, ab-interno | 24 | 29 | 0.86 |
| 31 | 2019 | Observational | Moderate or Advanced Glaucoma | Ab-interno | 21 | 21 | 0.67 |
| 32 | 2021 | Observational | Moderate or Advanced Glaucoma | Reinforced, ab-interno | 6 | 43 | 0.792 |
| 33 | 2021 | Observational | Moderate or Advanced Glaucoma | Ab-interno + MSICS | 12 | 343 | 0.843 |
| 34 | 2021 | Observational | Moderate or Advanced Glaucoma | Ab-interno | 12 | 53 | 0.528 |
| 35 | 2022 | Observational | Moderate or Advanced Glaucoma | Reinforced, ab-externo | 24 | 61 | 0.967 |
| 36 | 2023 | Case Control | Moderate or Advanced Glaucoma | Ab-externo + Goniotomy vs | 6 | 43 | 0.458 |

|  |  |  |  | Goniotomy |  |  |  |
| --- | --- | --- | --- | --- | --- | --- | --- |
| 37 | 2024 | Observational | Mild or Moderate Glaucoma | Reinforced, ab-interno + Phaco | 12 | 10 | 0.8 |
| 38 | 2024 | Observational | Mild or Moderate Glaucoma | Reinforced, ab-interno + Phaco | 12 | 117 | 0.81 |
| 39 | 2024 | Observational | Mild or Moderate Glaucoma | Reinforced, ab-interno + Phaco | 6 | 243 | 0.726 |
| 40 | 2025 | Observational | Mild or Moderate Glaucoma | Reinforced, ab-interno + Phaco | 30 | 24 | 0.74 |

**Figure 2.**
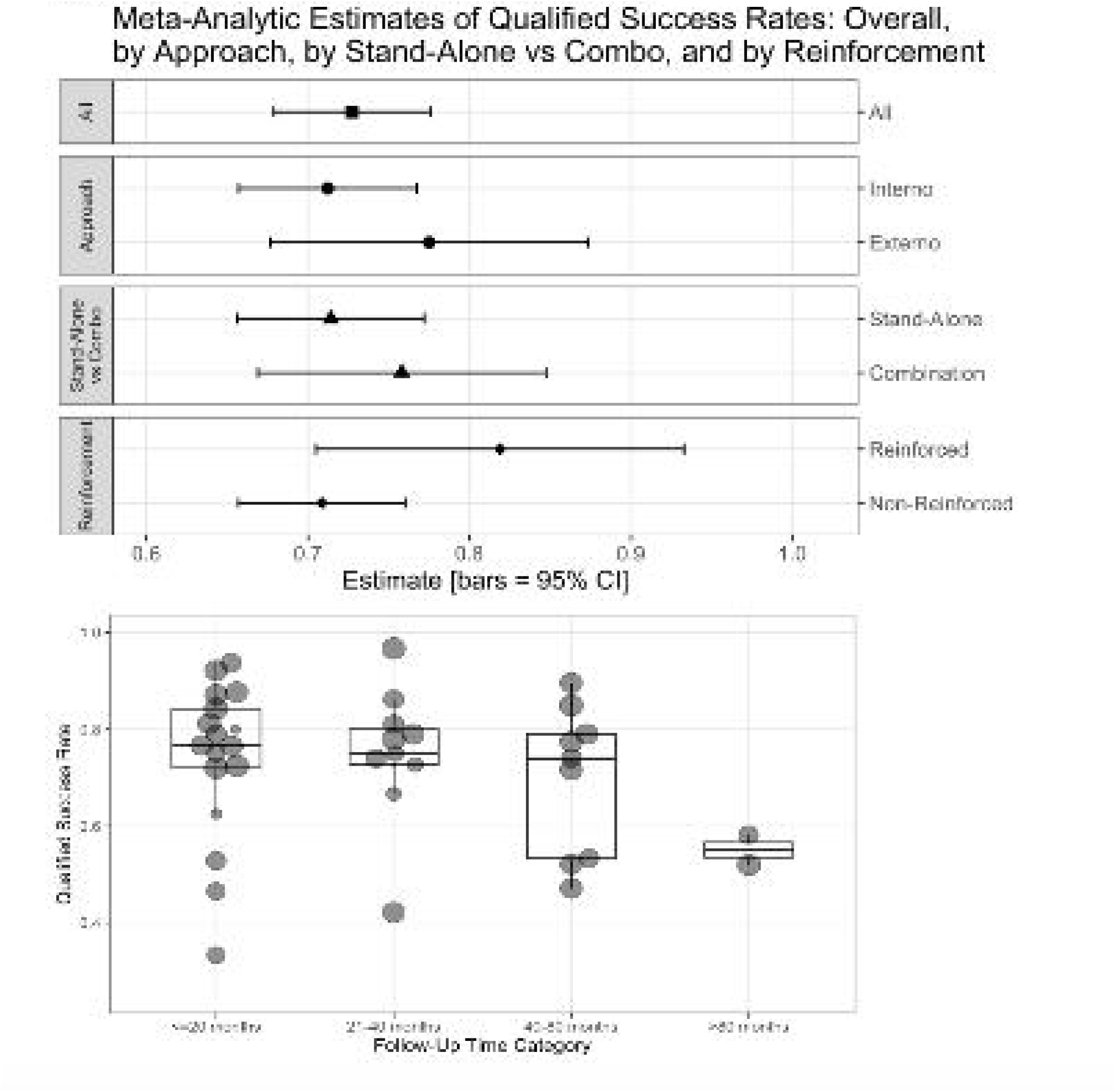
(a) Estimates of qualified success rates from meta-analyses. (b) Qualified success rates over time. Bars indicate 95% confidence intervals.

A final meta-analysis, with year published as a fixed effect, showed no trends in qualified success through time up to 80 months.

With respect to safety, there was no standardized reporting of surgical adverse events across studies, particularly in publications prior to 1970’s. Despite this lack of uniformity, the collective body of evidence consistently demonstrates a favorable safety profile for cyclodialysis. Across the studies, few patients experienced significant vision loss, even in cases involving ab-externo cyclodialysis procedures spanning up to 180 degrees. The recent studies of less invasive, ab-interno techniques report improved safety profile with the internal surgical approach.

Key adverse events reported in from the studies include transient hyphema 3%-79%, clinically significant hypotony 0.5-10.5%, persistent iritis 0-6%.

## DISCUSSION

We report the results of a systematic meta-analysis of surgical cyclodialysis treatment for the lowering of intraocular pressure in patients with glaucoma. The review spans more than a century of evolving surgical practice for the treatment of glaucoma across 40 peer-review clinical outcomes publications in over 4,000 subjects. It is the largest systematic meta-analysis review of a glaucoma interventional procedure to date.

With respect to the main outcome measure of qualified success, the overall efficacy of cyclodialysis exceeded 70%. This demonstrates evidence of clinical benefit in terms of IOP control achieved beyond the efficacy of topical pharmacologic therapy across the entire cohort of peer-review studies. Outcomes were similar regardless of procedure type, approach, or stand-alone versus combination procedures. The additional IOP lowering effect of a combined cataract procedure was demonstrable in our data set with 75.3% of the combined procedures vs 71.0% of the stand-alone cyclodialysis procedures achieving the primary success outcome. In the early years of cyclodialysis surgery, it was rarely performed in combination with intracapsular cataract extraction and later, with the advent of extracapsular cataract techniques it was combined with manual or phaco-emulsification cataract surgery. The higher level of IOP effect of combined cyclodialysis and cataract intervention is consistent with the experience of more recent glaucoma interventional studies which further substantiate the additive IOP-lowering efficacy of cataract extraction and glaucoma surgery. In addition, the data indicates a trend towards further IOP lowering (78.8% qualified success rate) when cyclodialysis is additionally reinforced with sub-scleral maintainers and spacers to prevent cleft restenosis and increase the durability and patency of the uveoscleral conduit.

To analyze for durability of effect, the dataset was segmented into different periods of post-operative outcomes- <20 months, 21-40 months, 40-80 months and> 80 months. While a slight attrition of IOP lowering effect can be seen over time from a peak of 74.6% at 20 months post-operative follow-up to 70.2% at 40-80 months, the differences among these post-operative time frames are not statistically significant. The are too few studies beyond 80 months of follow-up to provide an informative dataset for more extended follow-up. Similarly, there is not enough data in the early post-operative period before 6 months to explore if there is any temporal relationship to the post-surgical remodeling and healing of the cyclodialysis. Regardless, the long-term effect across all studies is consistent and sustained. Also, reinforced cyclodialysis procedures, while demonstrating higher efficacy in terms of the primary outcome over 12 and 24 months, did not have data to determine efficacy beyond the 5-year follow-up.

Our analysis also demonstrates comparable efficacy and safety between ab-externo and ab-interno surgical approaches. In fact, a trend towards slightly better IOP lowering effect seems to transpire from the ab-interno studies (77.5% vs 71.2%, respectively). This is clinically important as the size of the cyclodialysis cleft in the ab-externo procedures is generally much larger than the one performed ab-interno, particularly with the more recent interventional techniques, where cleft sizes are discrete and minimally invasive within 500-1,000 microns. This may substantiate the clinical paradigm of the continuity of the uveoscleral outflow where a single discrete entry point can tap into the absorptive outflow capacity of the entire suprachoroidal space. It may also indicate better healing and less fibrosis with minimal intervention and less surgical trauma. The benefits of this more interventional ab-interno approach are also seen in terms of safety where a slightly lower rate of key ocular adverse events are reported in an otherwise homogeneous and favorable safety profile of the cyclodialysis procedure. The overall safety of the procedure is characterized by a relatively low incidence of serious or sight-threatening adverse events and appears more comparable to the newer interventional glaucoma procedures. The characteristic complications of hypotony maculopathy, bleb leaks or loss of visual acuity often reported with trans-scleral glaucoma filtration procedures such as trabeculectomy and external shunt drainage implants are not seen as a major concern with cyclodialysis where aqueous outflow enhancement remains internal to the uveoscleral pathway, there is no violation of the scleral wall and there is no exogenous implantable hardware ^27,28^.

There are several limitations of our analysis. A key limitation is that all 40 available cyclodialysis studies were non-randomized, observational real-world clinical reports. While this is informative of real-world effectiveness, there are confounding factors such as lack of medication wash-out, no protocolized medication re-introduction, no standardized definition of efficacy outcomes, lack of standardized indications for the surgery, and lack of treatment controls. Most studies also had poor definition of mild, moderate or severe glaucoma subtypes and primary open angle versus secondary open angle glaucoma as they did not consistently provide disease category definitions nor baseline visual field data. Another limitation is the insufficient understanding of the precise anatomical location, structural dimensions, and composition of cyclodialysis clefts—possibly critical factors influencing their efficacy and consistency. It is well established that both trabecular and uveoscleral outflows are segmental, making the exact site and scale of intervention potentially useful information ^29-34^. However, few studies provide detailed accounts of surgical techniques with definition of cleft size nor do they incorporate companion imaging to assess cleft size over time. Furthermore, lack of imaging techniques especially in earlier studies prevented detection of clinically inapparent cystoid macular edema or hypotony maculopathy. Finally, improvements in medical treatment over the last 100+ years would likely favor qualified success rates in more recent studies. Nevertheless, given the span in clinical evidence over 125 years of evolving surgical practice since leopold Heine first introduced the procedure^35^, this integrated evidence-based summary of rather heterogenic reports and data, provides surprisingly consistent evidence of IOP lowering effect and risk-benefit profile of uveoscleral outflow intervention. These observations may be useful in informing and encouraging future surgical innovations in the anterior suprachoroidal space.

## CONCLUSION

In conclusion, cyclodialysis represents an established, clinically effective surgical option for glaucoma management, characterized by significant IOP lowering and a favorable safety profile. Newer approaches for reinforcement of the cleft could further enhance the durability of the uveoscleral conduit over the long term. As surgical techniques and instrumentation continue to evolve with advanced interventional and bio-interventional approaches, cyclodialysis-based procedures offer a non-pharmacologic enhancement of the uveoscleral outflow pathway in the management of glaucoma.

## Data Availability

All data produced in the present study are available upon reasonable request to the authors

## ADDENDUM

References of studies included in the analysis

| Study No. | Year Pub. | Reference |
| --- | --- | --- |
| 1 | 1920 | M . f. Augenh. 1920 , v64 p 433 Salus , R. Die Zyklodialyse Klin “Die Zyklodialyse” by R. Salus, published in 1920 in Klinische Monatsblätter für Augenheilkunde (Clinical Monthly Reports for Ophthalmology), Volume 64, pages 433–509. Zykloskopische Untersuchungen über das Verhalten des Strahlenkörpers nach derHeinischen Operation |
| 2 | 1920 | M . f. Augenh. 1920 , v64 p 433 Salus , R. Die Zyklodialyse Klin “Die Zyklodialyse” by R. Salus, published in 1920 in Klinische Monatsblätter für Augenheilkunde (Clinical Monthly Reports for Ophthalmology), Volume 64, pages 433–509. Zykloskopische Untersuchungen über das Verhalten des Strahlenkörpers nach derHeinischen Operation |
| 3 | 1930 | Archiv für Augenheilkunde (Archive for Ophthalmology), 1930, Volume 102, pages 626–655. Stein, R Die Dauerresultate der Zyklodialyse |
| 4 | 1931 | Arch Ophthalmol1931;5(3):327-333. doi:10.1001. |
| 5 | 1937 | Ztschr. f. Augenh. 92:139,1937 Hausmann, G.: Zur has der Dauerrsul- tate der Zyklodialyse |
| 6 | 1940 | Archives of ophthalmology (1879), 1940-02, Vol.23 (2), p.270-300 |
| 7 | 1941 | Arch of Ophth 1941 v25 Apr p 674 Sugar. H.E. Gonioscopy and Glaucoma |
| 8 | 1946 | Acta Ophthalmol, 24 (1946), p. 27. Louhala, T., and Teraskel,H.: On the Results of Different Glaucoma Operations |
| 9 | 1946 | Am J Ophthalmol 1946 Jul;29:848-53 McPherson , S. D. Cyclodialysis in the treatment of glaucoma |
| 10 | 1947 | Am J Ophthalmol 1947 Sep;30(9):1063-73 |
| 11 | 1947 | Am. J. Ophth. 30:451, 1947. Sugar, H. S.: Cyclodialysis: A Follow-up Study |
| 12 | 1947 | Calif Med 1947 Aug;67(2):78-83 |
| 13 | 1949 | Arch Ophthal 1949 Nov;42(5):606-19 |
| 14 | 1949 | Arch. Ophth. 42:606, 1949 |
| 15 | 1954 | American journal of ophthalmology, 1954-11, Vol.38 |
| 16 | 1957 | Acta ophthalmologica (Oxford, England), 1957-10, Vol.35 (5), p.528-542 |
| 17 | 1957 | Acta ophthalmologica (Oxford, England), 1957-10, Vol.35 (5), p.528-542 |
| 18 | 1958 | Arch Ophthalmol. 1958; 59(4):507-514 |
| 19 | 1958 | Trans Am Ophthalmol Soc. 1958;56:263–274. |
| 20 | 1960 | Arch Ophthalmol. 1960;64(3):408-412. |
| 21 | 1969 | American Journal of Ophthalmology, Volume 67, Issue 4, 522 – 526 |
| 22 | 1976 | Ophthalmic Surg 1976 Summer;7(2):62-73 |
| 23 | 1986 | Acta Ophthalmol (Copenh) 1986 Apr;64(2):142-5 |
| 24 | 1986 | Acta Ophthalmol 64:142–145. Hansen S, Laursen AB (1986) Visualized cyclodialysis-an additional option in glaucoma surgery. |
| 25 | 1995 | Ophthalmologe 1995 Aug;92(4):531-5 |
| 26 | 1998 | Ophthalmic Surg Lasers 1998 Dec;29(12):962-8 |
| 27 | 2013 | Ophthalmic Surgery, Lasers and Imaging Retina, 2013;12(2):92-94 |
| 28 | 2013 | Ophthalmic Surgery, Lasers and Imaging Retina, 2013;29(12):962-968 |
| 29 | 2015 | J Glaucoma 2015 Jan;24(1):37-44. |
| 30 | 2019 | Medicina v Kuzbasse, 18, 20190201, 61; ISSN 1819-0901 |
| 31 | 2019 | Medicina v Kuzbasse, 18, 20190201, 61; ISSN 1819-0901 |
| 32 | 2021 | ISSN 1816-5095 (print); ISSN 2500-0845 (online) https://doi.org/10.18008/1816-5095-2021-1-61-69 |
| 33 | 2021 | Papa-Vettorazzi, Mario R., et al. "Results of combined surgery, mSICS and cyclodialysis, in patients with glaucoma and cataract. Real-life experience, efficacy and safety Resultados de cirugía combinada, mSICS y ciclodíálisis, en pacientes con glaucoma y |
| 34 | 2021 | Papa-Vettorazzi, Mario R., et al. "Results of combined surgery, mSICS and cyclodialysis, in patients with glaucoma and cataract. Real-life experience, efficacy and safety Resultados de cirugía combinada, mSICS y ciclodíálisis, en pacientes con glaucoma y |
| 35 | 2022 | Kumar, V., Shradqa, A., Frolov, M., Dushina, G. and Bezzabotnov, A. (2022), Acta Ophthalmol, 100:.. https://doi.org/10.1111/j.1755-3768.2022.081 |
| 36 | 2023 | Graefes Arch Clin Exp Ophthalmol 2023 Oct;261(10) :2917-2925 |
| 37 | 2024 | Br J Ophthalmol 2024 Jan 29;108(2):217-222 |
| 38 | 2024 | Clin Ophthalmol 2024 Dec 6;18:3605-3614 |
| 39 | 2024 | J Clin Med 2024 Aug 6;13(16):4593 |
| 40 | 2025 | Ophthalmology Science 2025 Feb (In Press) |

